# A national retrospective analysis of the management of patients presenting as an emergency with a foreign body in the oesophagus

**DOI:** 10.64898/2025.11.30.25341290

**Authors:** Rohan Aggarwal, Dominic King, Ben Coupland, David McNulty, Amar Srinivasa, Hasan Haboubi, Caroline Turnbull, Riadh Jazrawi, Nigel Trudgill

## Abstract

**Background and Aims:** *Emergency a*dmission with foreign body in the oesophagus (FBO) commonly requires endoscopic removal. Oesophageal food bolus obstruction is often due to eosinophilic oesophagitis (EoE). Patients with food bolus obstruction should undergo endoscopy and multilevel oesophageal biopsies to exclude EoE. This national retrospective study examined the management, including endoscopy and follow-up, of patients presenting as an emergency with FBO.

**Methods:** *The Hospita*l Episode Statistics database was used to identify patients over 18 with FBO presenting as an emergency in England using the ICD-10 code T18.1 between 2008 and 2019. Logistic regression analysis was used to assess factors associated with undergoing endoscopy and biopsy.

**Results:** 27,063 patients were identified: 65.5% male; median age 57(IQR 41-73) years; and 47.2% were admitted under Ear, Nose & Throat (ENT). 75.7% underwent endoscopy (94% within a week of admission) but only 19.8% had biopsies taken within 6 months of admission. 0.4% were coded with a perforation related to endoscopy for FBO. 70% of ENT patients underwent endoscopy but only 11.9% had biopsy to exclude EOE, compared with 83% of patient admitted under General Medicine undergoing endoscopy and 29.5% biopsy. Endoscopy and biopsy was associated with: older age (e.g. 61-70 OR 1.42 (95% CI 1.26-1.61)), males (females 0.67(0.62-0.72)), the least deprived (1.25 (1.13-1.38)), later diagnosis year (2019 1.42 (1.21-1.66)), and admission under General Medicine (2.68(2.48-2.88)) or Gastroenterology (3.03(2.61-3.51)) but not with NHS trust FBO volume. 33.4% received relevant outpatient follow-up within 12 months of FBO admission: 26% of patients admitted under General Medicine were referred to gastroenterology for follow-up, but only 12.1% of those admitted under ENT.

**Conclusions:** 75.7% of patients presenting with FBO undergo endoscopy but few (19.8%) had biopsies taken to exclude this common presentation of EOE. Pathways for the management of food bolus obstruction require re-design and unless the airway is impaired, this condition should be managed under General Medicine.

## Introduction

Foreign body in the oesophagus is a common emergency presentation in both children and adults. Foreign bodies range from in-organic materials, e.g. small batteries, to food boluses or bones^1^. Among adults, food bolus obstruction makes up the vast majority cases of foreign body in the oesophagus and has an estimated annual incidence rate of 13 per 100,000 of the general population, which has been increasing over the last decade and a half^2,3^. Eosinophillic oesophagitis (EoE) is an immune-allergic condition that affects the oesophagus and is a common underlying cause of food bolus obstruction^4,5^. EoE is diagnosed through a combination of clinical features, endoscopic findings, and oesophageal histology^6^. EOE more commonly affects men with a male: female ratio of 3:1, the incidence in adults peaks between the ages of 30 and 40 and its incidence and prevalence are increasing^7^. Endoscopic intervention to treat food bolus obstruction represents an opportunity to make a diagnosis of EOE and prevent future emergency episodes through effective EoE therapy^8^.

Macroscopic findings on endoscopy such as oesophageal mucosal trachealisation, oesophageal furrows or rings can aid in recognition of EoE, but the appearance of the oesophagus can be normal^9^. To reliably diagnose EoE, a minimum of six oesophageal biopsies should be taken from two or more levels of the oesophagus, focussing on areas with endoscopic mucosal abnormalities^10,11^.

The aim of this large national retrospective study was to investigate the management of patients presenting to hospital as an emergency with foreign body in the oesophagus and in particular whether patients underwent endoscopy and biopsy to exclude potential underlying EOE.

## Methods and Materials

### Data source

Hospital Episode Statistics (HES) is a database managed by NHS Digital which includes information on all episodes of National Health Service (NHS) secondary care treatment within England. HES contains diagnostic (International Classification of Diseases version 10 (ICD10)) and procedural (Office of Population Censuses and Surveys Classification of Interventions and Procedures 4th revision (OPCS-4)) data. Demographic and geographic data for each patient are also recorded. Individuals can be tracked through their hospital admissions via a unique identifier. The HES data sharing agreement prohibits the publication of potentially identifiable data and it is for this reason that patient counts of five or less are suppressed from publication.

### Inclusion criteria

All patients aged over 18 with foreign body in the oesophagus were identified in HES by using the code ICD-10-T18.1 between the dates 01/01/2008 and 31/12/2019. OPCS-4 codes for fibreoptic and rigid endoscopic examination and biopsy of the oesophagus were used to identify patients who underwent endoscopy and/or biopsy within a 6-month period following an emergency presentation with foreign body in the oesophagus.

### Exclusion criteria

Patients were excluded if they had missing or invalid age, sex, deprivation quintile, or region of residence data or were non-resident in England. Patients with a diagnosis of upper gastrointestinal cancer were excluded, as well as patients diagnosed with foreign body in the oesophagus during a previous hospital admission.

### Data validation

To assess the validity of the diagnostic and procedural coding for foreign body in the oesophagus, a list of patients meeting the same ICD-10 and OPCS-4 coding criteria was provided by the local coding department at Sandwell & West Birmingham NHS Trust. The accuracy of the coding was then assessed by consulting the electronic patient records to establish the nature of the foreign body in the oesophagus. To further validate the coding accuracy, data on endoscopic extraction of foreign bodies in the oesophagus for the whole of the UK for four years was examined in the National Endoscopy Database.

### Demographic data

Patient age, sex, deprivation status and ethnicity were identified from emergency admission coding. Age was divided into quartiles 18-30, 31-40, 41-50, 51-60, 61-70, 71-80 and over 80. Ethnicity was stratified into white, Asian, other minority ethnicities and unknown. The Charlson comorbidity index, a measure of multimorbidity in patients and previously validated in HES, was calculated using secondary diagnostic coding^12^. Deprivation quintiles were calculated from the Index of Multiple Deprivations, a classification based on income, employment, crime and living environment^13^. Deprivation quintile 5 is the least deprived quintile, while quintile 1 is the most deprived.

### Outcome measures

The primary outcome measure in this study was the number of patients presenting as an emergency with foreign body in the oesophagus who underwent endoscopy with biopsy within 6 months of presentation to hospital. A secondary outcome measure was the number of patients who attended follow-up as an outpatient with gastroenterology within 12 months of their emergency presentation.

### Proposed algorithm for management of food bolus obstruction

Based on study data, a proposed algorithm for inpatient management of food bolus obstruction was then developed.

### Statistical analysis

Demographic data are presented as number and percentage where applicable. Age is presented as median and interquartile range (IQR). Logistic regression analysis models were constructed to examine associations with undergoing endoscopy and endoscopy with biopsy with estimates presented as odds ratios (OR). Variables included in the models were sex, age group, ethnicity, comorbidity score category, deprivation quintile, diagnosis year, provider volume of foreign body in the oesophagus admissions and admitting speciality for the emergency admission (Ear Nose and Throat (ENT), Accident & Emergency, Gastroenterology, General Medicine, General Surgery). P values of <0.05 were considered statistically significant. All statistical analysis was carried out using Stata version 15.

### Ethics

HES data are available under a data sharing agreement with NHS Digital for the purpose of service evaluation. Ethics approval is therefore not required. The study was registered in the Audit and Clinical Effectiveness Department at Sandwell & West Birmingham NHS Trust and the Department of Informatics at University Hospitals Birmingham NHS Foundation Trust.

## Results

### Validation data

All admissions aged over 18 years to Sandwell & West Birmingham NHS Trust with code T18.1 for ‘foreign body in the oesophagus’ were examined between July 2014 and June 2020. 107 patients were identified and food boluses were the cause for the foreign body in the oesophagus in 81% of this cohort.

In the National Endoscopy Database between January 2018 and December 2021, 1230 patients underwent endoscopy to remove a foreign body from the oesophagus. In 1124 patients (91%) a food bolus was removed and in 106 patients (9%) a foreign body other than food was removed.

### Cohort characteristics

Between 1^st^ Jan 2008 and 31^st^ Dec 2019, 30,487 patients with an emergency admission for foreign body in the oesophagus met the inclusion criteria. 1831 were excluded for having missing or invalid details and 154 due to a prior diagnosis of foreign body in the oesophagus. A further 1,439 were excluded due to a prior upper GI cancer diagnosis. A final cohort of 27,063 patients was included in the study (Figure 1).

**Figure 1.**
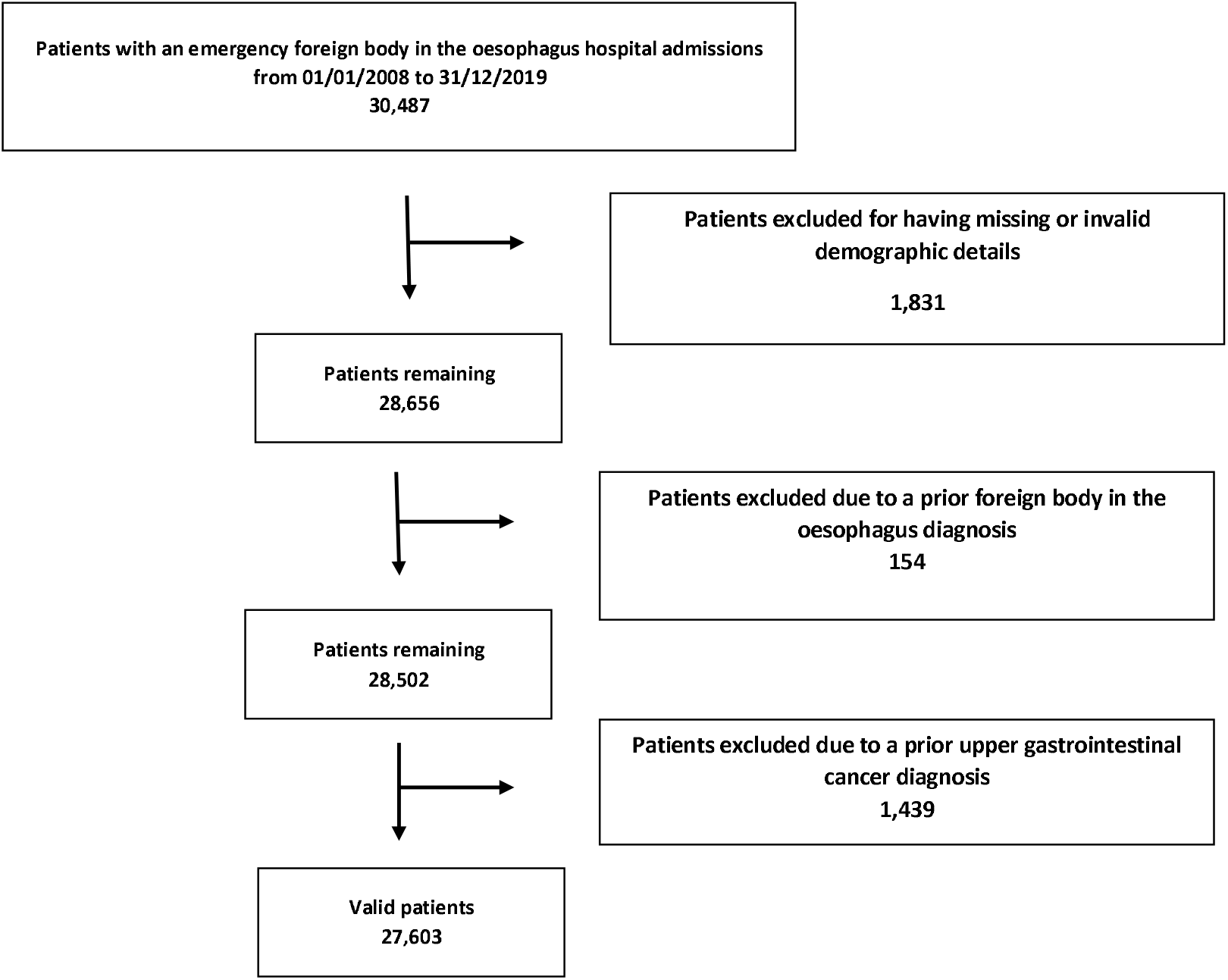
Study flow chart.

The characteristics of the patients studied are shown in Table 1. 66% of the population were male and the median age was 57 (IQR 41-73) years. 47.2% of patients were admitted under ENT, 33.6% under General Medicine, 9.1% under General Surgery, 6.5% under Accident and Emergency and 3.6% under Gastroenterology. This pattern changed over the study period with 64% of patients with foreign body in the oesophagus admitted under ENT in 2008 but only 33% in 2019, with General Medicine admitting 17% in 2008 and 48% of such patients in 2019.

**Table 1.**
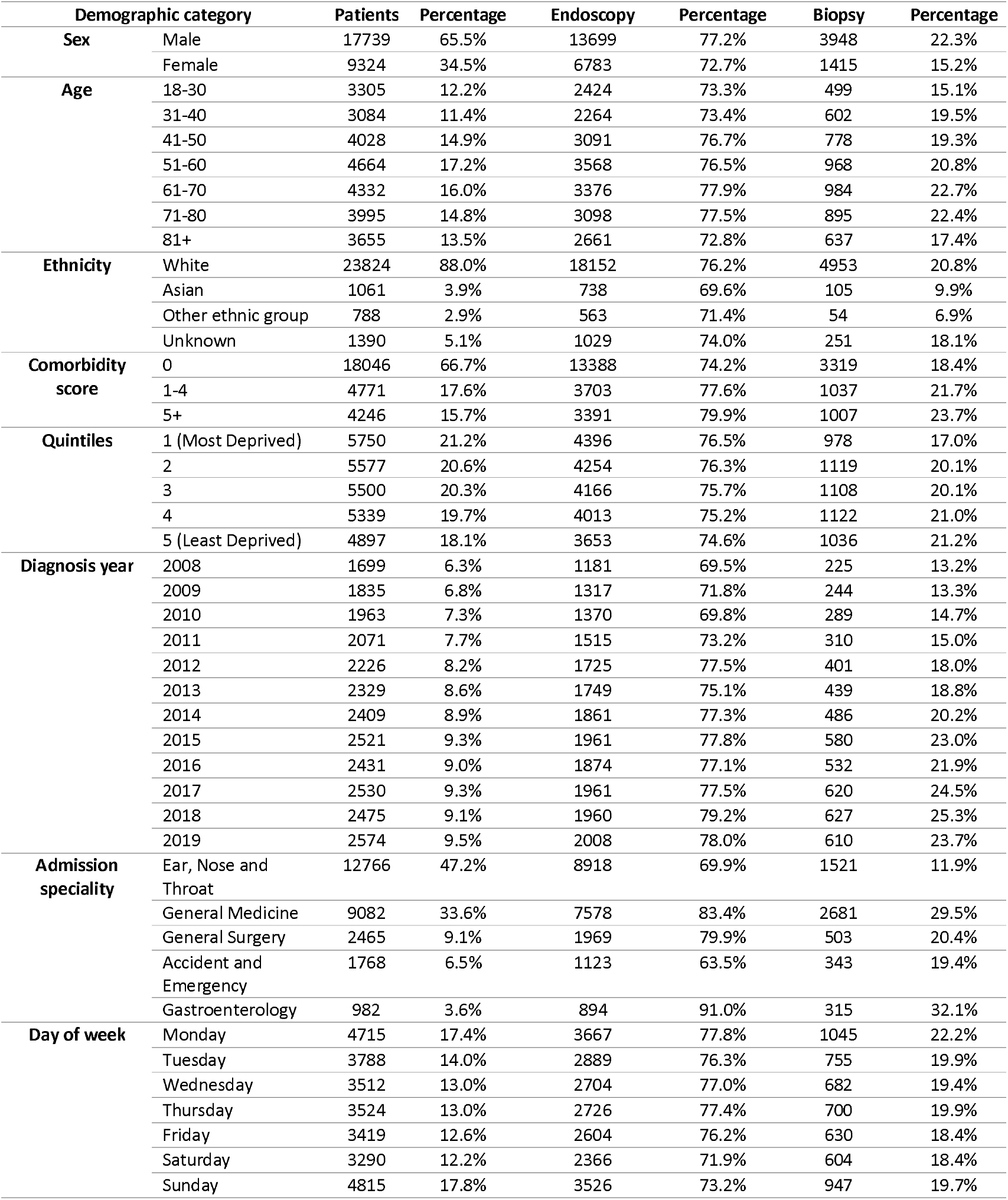
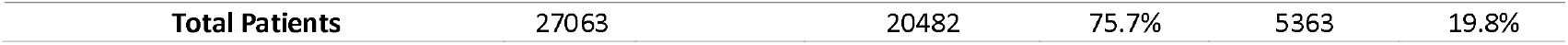
Demographic data of study patients including those undergoing endoscopy with or without oesophageal biopsy within 6 months of emergency admission with foreign body in the oesophagus.

The annual number of admissions for foreign body in the oesophagus increased initially during the study period from 1699 in 2008 to 2521 in 2015 but then has remained relatively static. The percentage of patients who underwent endoscopy within 6 months of admission also increased from 69.5% in 2008 to 79.2% in 2018 (p<0.001). The percentage of endoscopies with biopsies increased from 13.2% in 2008 to 25.3% in 2018 (p<0.001).

Out of 20,482 endoscopies carried out within 6 months of admission, 82.5% were performed using a flexible endoscope and endoscopies carried out with flexible endoscopes accounted for 99.4% of endoscopies with biopsies.

Patients were more likely to be admitted with foreign body in the oesophagus on a Monday (17.4% of total) or a Sunday (17.8% of total), compared with other days of the week (Monday and Sunday versus all other days by one way analysis of variance p<0.001). The number of patients undergoing endoscopy with biopsy was higher on a Monday (22.2%) compared to the rest of the week (18.4%-19.9%) (p<0.001). There was no evidence of seasonality affecting patients presenting with foreign body in the oesophagus.

### Endoscopy and biopsy for foreign body in the oesophagus

Overall, 75.7% patients underwent endoscopy within 6 months of their emergency presentation and 19.8% had biopsies taken. 78% of the endoscopies and 43.8% of the total biopsies were performed within one week of their emergency presentation with foreign body in the oesophagus.

69.9% of patients admitted under ENT underwent endoscopy compared to 83.4% of those admitted under General Medicine and 91% admitted under Gastroenterology. Biopsies were carried out in 11.9% of patients admitted under ENT, compared to 29.5% of patients admitted under General Medicine and 32.1% under Gastroenterology. The percentage of patients undergoing endoscopy and biopsy under ENT increased over the study period (8.5% in 2008 and 11.7% in 2019, p=0.01)) but increased more under General Medicine (23% in 2008 to 31.7% in 2019, p=0.001).

Over the study period, there were 319 endoscopic dilatations (0.9% of all endoscopies) in patients with foreign body in the oesophagus. 99 patients (0.4% of all endoscopies) undergoing endoscopy for foreign body in the oesophagus were coded as having a perforation. Of the 319 patients who had a dilatation performed at their endoscopy, only 37 (11.6%) had a biopsy coded at some point in the six months after their foreign body in the oesophagus emergency admission.

### Logistical regression analysis of factors associated with endoscopy and biopsy following a foreign body in the oesophagus emergency admission

Tables 2 and 3 show the results of logistic regression analysis of associations with undergoing endoscopy and separately endoscopy with biopsy. Factors associated with undergoing endoscopy included: age – 41-50 years (odds ratio 1.16 (95%CI 1.04-1.29)) and 61-70 (1.14(1.02-1.27) compared with 18-30; increasing comorbidity - 1.20 (1.10-1.32); later diagnosis year - 1.27 (1.10-1.45) in 2019; admission under - General Medicine (2.02 (1.88-2.17)), Gastroenterology (4.07 (3.26-5.09)) or General Surgery (1.67 (1.50-1.86)) compared with ENT; and admission on any day of the week other than a Saturday or Sunday. Factors associated with being less likely to undergo endoscopy were: age over 81 (0.82 (0.73-0.92)) compared with 18-30; female - 0.83 (0.78-0.88); less deprivation – quintile 5 0.88 (0.80-0.97); Asian ethnicity - 0.86 (0.74-0.98); and admission under Accident and Emergency - 0.73 (0.66-0.81).

**Table 2.** Logistic regression analysis of factors associated with endoscopy within 6 months of a foreign body in the oesophagus emergency admission.

|  |  | Odds ratio | P Value | 95% CI |  |
| --- | --- | --- | --- | --- | --- |
| **Sex** | Male (baseline) |  |  |  |  |
|  | Female | 0.83 | **<0.001** | 0.78 | 0.88 |
| **Age** | 18-30 (baseline) |  |  |  |  |
|  | 31-40 | 0.98 | 0.775 | 0.88 | 1.10 |
|  | 41-50 | 1.16 | **0.008** | 1.04 | 1.29 |
|  | 51-60 | 1.11 | 0.058 | 1.00 | 1.23 |
|  | 61-70 | 1.14 | **0.020** | 1.02 | 1.27 |
|  | 71-80 | 1.07 | 0.247 | 0.95 | 1.20 |
|  | 81+ | 0.82 | **0.001** | 0.73 | 0.92 |
| **Ethnicity** | White (baseline) |  |  |  |  |
|  | Asian | 0.85 | **0.025** | 0.74 | 0.98 |
|  | Other ethnic group | 0.93 | 0.353 | 0.79 | 1.09 |
|  | Unknown | 0.92 | 0.214 | 0.81 | 1.05 |
| **Comorbidity score** | 0 (baseline) |  |  |  |  |
|  | 1-4 | 1.15 | **0.001** | 1.06 | 1.24 |
|  | 5+ | 1.21 | **<0.001** | 1.10 | 1.32 |
| **Deprivation quintile** | 1 (Most Deprived) (baseline) |  |  |  |  |
|  | 2 | 0.97 | 0.508 | 0.89 | 1.06 |
|  | 3 | 0.93 | 0.113 | 0.85 | 1.02 |
|  | 4 | 0.91 | **0.044** | 0.83 | 1.00 |
|  | 5 (Least Deprived) | 0.88 | **0.007** | 0.80 | 0.97 |
| **Diagnosis year** | 2008 | 1.03 | 0.729 | 0.89 | 1.18 |
|  | 2009 | 1.14 | 0.078 | 0.99 | 1.31 |
|  | 2010 (baseline) |  |  |  |  |
|  | 2011 | 1.16 | **0.034** | 1.01 | 1.34 |
|  | 2012 | 1.43 | **<0.001** | 1.24 | 1.65 |
|  | 2013 | 1.23 | **0.003** | 1.07 | 1.41 |
|  | 2014 | 1.35 | **<0.001** | 1.18 | 1.55 |
|  | 2015 | 1.36 | **<0.001** | 1.19 | 1.57 |
|  | 2016 | 1.28 | **<0.001** | 1.11 | 1.47 |
|  | 2017 | 1.29 | **<0.001** | 1.12 | 1.48 |
|  | 2018 | 1.37 | **<0.001** | 1.19 | 1.58 |
|  | 2019 | 1.27 | **0.001** | 1.10 | 1.45 |
| **Trust volume tertile** | Low (baseline) |  |  |  |  |
|  | Medium | 0.88 | **0.035** | 0.78 | 0.99 |
|  | High | 0.89 | 0.055 | 0.80 | 1.00 |
| **Admission speciality** | Ear, Nose and Throat (baseline) |  |  |  |  |
|  | General Medicine | 2.03 | **<0.001** | 1.90 | 2.19 |
|  | General Surgery | 1.67 | **<0.001** | 1.50 | 1.86 |
|  | Accident and Emergency | 0.73 | **<0.001** | 0.66 | 0.81 |
|  | Gastroenterology | 4.11 | **<0.001** | 3.28 | 5.14 |
| **Day of the week** | Sunday (baseline) |  |  |  |  |
|  | Monday | 1.21 | **<0.001** | 1.10 | 1.33 |
|  | Tuesday | 1.14 | **0.01** | 1.03 | 1.26 |
|  | Wednesday | 1.18 | **0.002** | 1.06 | 1.31 |
|  | Thursday | 1.21 | **<0.001** | 1.09 | 1.34 |
|  | Friday | 1.13 | **0.02** | 1.02 | 1.25 |
|  | Saturday | 0.92 | 0.13 | 0.84 | 1.02 |

**Table 3.**
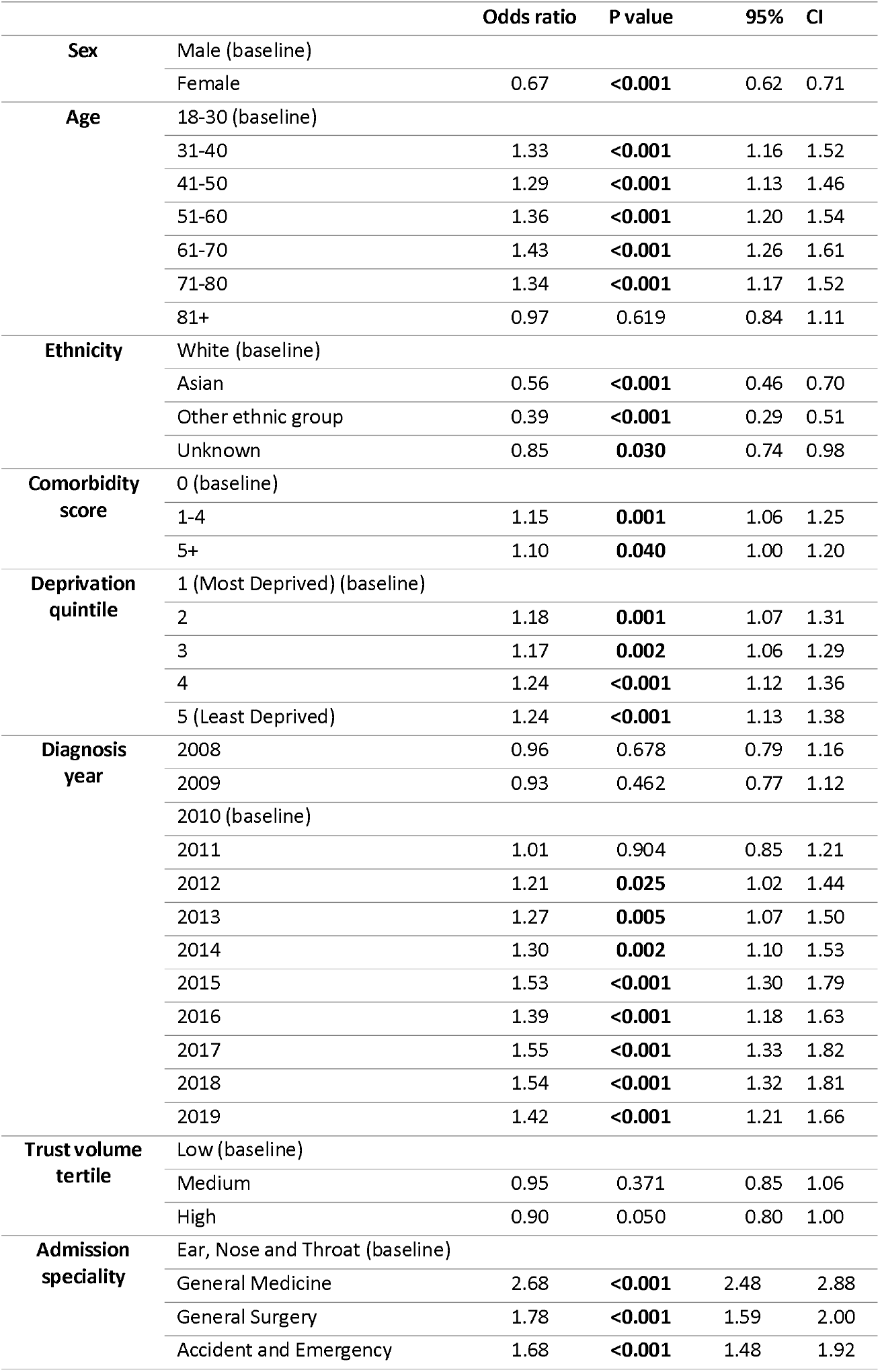
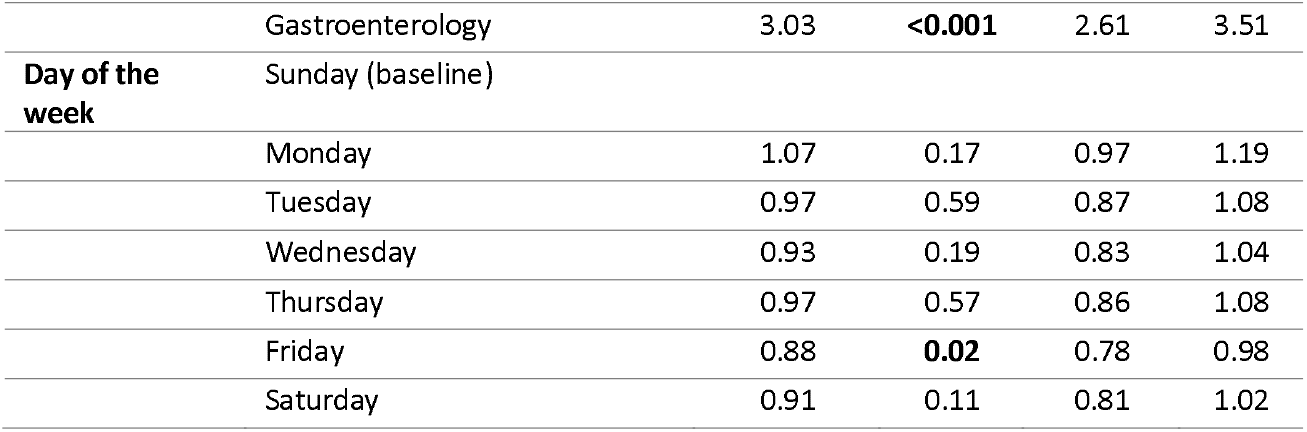
Logistic regression analysis of factors associated with endoscopy with biopsy within 6 months of foreign body in the oesophagus emergency admission.

|  |  | Odds ratio | P value | 95% CI |
| --- | --- | --- | --- | --- |
| **Sex** | Male (baseline) |  |  |  |
|  | Female | 0.67 | <0.001 | 0.62 0.71 |
| **Age** | 18-30 (baseline) |  |  |  |
|  | 31-40 | 1.33 | <0.001 | 1.16 1.52 |
|  | 41-50 | 1.29 | <0.001 | 1.13 1.46 |
|  | 51-60 | 1.36 | <0.001 | 1.20 1.54 |
|  | 61-70 | 1.43 | <0.001 | 1.26 1.61 |
|  | 71-80 | 1.34 | <0.001 | 1.17 1.52 |
|  | 81+ | 0.97 | 0.619 | 0.84 1.11 |
| **Ethnicity** | White (baseline) |  |  |  |
|  | Asian | 0.56 | <0.001 | 0.46 0.70 |
|  | Other ethnic group | 0.39 | <0.001 | 0.29 0.51 |
|  | Unknown | 0.85 | 0.030 | 0.74 0.98 |
| **Comorbidity score** | 0 (baseline) |  |  |  |
|  | 1-4 | 1.15 | 0.001 | 1.06 1.25 |
|  | 5+ | 1.10 | 0.040 | 1.00 1.20 |
| **Deprivation quintile** | 1 (Most Deprived) (baseline) |  |  |  |
|  | 2 | 1.18 | 0.001 | 1.07 1.31 |
|  | 3 | 1.17 | 0.002 | 1.06 1.29 |
|  | 4 | 1.24 | <0.001 | 1.12 1.36 |
|  | 5 (Least Deprived) | 1.24 | <0.001 | 1.13 1.38 |
| **Diagnosis year** | 2008 | 0.96 | 0.678 | 0.79 1.16 |
|  | 2009 | 0.93 | 0.462 | 0.77 1.12 |
|  | 2010 (baseline) |  |  |  |
|  | 2011 | 1.01 | 0.904 | 0.85 1.21 |
|  | 2012 | 1.21 | 0.025 | 1.02 1.44 |
|  | 2013 | 1.27 | 0.005 | 1.07 1.50 |
|  | 2014 | 1.30 | 0.002 | 1.10 1.53 |
|  | 2015 | 1.53 | <0.001 | 1.30 1.79 |
|  | 2016 | 1.39 | <0.001 | 1.18 1.63 |
|  | 2017 | 1.55 | <0.001 | 1.33 1.82 |
|  | 2018 | 1.54 | <0.001 | 1.32 1.81 |
|  | 2019 | 1.42 | <0.001 | 1.21 1.66 |
| **Trust volume tertile** | Low (baseline) |  |  |  |
|  | Medium | 0.95 | 0.371 | 0.85 1.06 |
|  | High | 0.90 | 0.050 | 0.80 1.00 |
| **Admission speciality** | Ear, Nose and Throat (baseline) |  |  |  |
|  | General Medicine | 2.68 | <0.001 | 2.48 2.88 |
|  | General Surgery | 1.78 | <0.001 | 1.59 2.00 |
|  | Accident and Emergency | 1.68 | <0.001 | 1.48 1.92 |

|  |  |  |  |  |  |
| --- | --- | --- | --- | --- | --- |
|  | Gastroenterology | 3.03 | **<0.001** | 2.61 | 3.51 |
| **Day of the week** | Sunday (baseline) |  |  |  |  |
|  | Monday | 1.07 | 0.17 | 0.97 | 1.19 |
|  | Tuesday | 0.97 | 0.59 | 0.87 | 1.08 |
|  | Wednesday | 0.93 | 0.19 | 0.83 | 1.04 |
|  | Thursday | 0.97 | 0.57 | 0.86 | 1.08 |
|  | Friday | 0.88 | **0.02** | 0.78 | 0.98 |
|  | Saturday | 0.91 | 0.11 | 0.81 | 1.02 |

Factors associated with undergoing endoscopy with biopsy included: increasing age – e.g. 31-40 years 1.33 (1.16-1.52) compared with 18-30; less deprivation - 1.25 (1.13-1.38); later diagnosis year 2019 1.42 (1.21-1.67); increasing co-morbidity; and admission under any speciality other than ENT – e.g. General Medicine 2.68 (2.48-2.88). Factors associated with being less likely to undergo endoscopy with biopsy were: female - 0.67 (0.62-0.71); and any minority ethnicity.

### Outpatient assessment following an emergency admission with a foreign body in the oesophagus

Patients admitted with food bolus obstruction should be reviewed in gastroenterology outpatients following discharge for further assessment and management to prevent recurrent episodes^14^. The outpatient follow-up organised either with the admitting speciality or with Gastroenterology in the 12 months following an emergency presentation with foreign body in the oesophagus are shown in Table 4. 33.4% received a follow-up outpatient appointment within 12 months of an admission with foreign body in the oesophagus. 21% received a follow-up appointment with the admitting speciality and 18.3% with gastroenterology.

**Table 4.** Outpatient follow-up of patients in the 12 months following a foreign body in the oesophagus admission by admitting specialty.

|  | Overall cohort |  | Follow up with same speciality |  |  |  |  |  | Follow up with Gastroenterology |  |  |  |  |  | Overall outpatient numbers |  |
| --- | --- | --- | --- | --- | --- | --- | --- | --- | --- | --- | --- | --- | --- | --- | --- | --- |
|  | Patients | Share | 0-6 months |  | 6-12 months |  | Overall |  | 0-6 months |  | 6-12 months |  | Overall |  | Patients | Share |
| Admission speciality |  |  | Patients | Share | Patients | Share | Patients | Share | Patients | Share | Patients | Share | Patients | Share |  |  |
| Ear, Nose and Throat | 12766 | 47.2% | 2957 | 23.2% | 823 | 6.4% | 3228 | 25.3% | 1164 | 9.1% | 812 | 6.4% | 1544 | 12.1% | 4351 | 34.1% |
| General Medicine | 9082 | 33.6% | 1115 | 12.3% | 692 | 7.6% | 1413 | 15.6% | 1974 | 21.7% | 1081 | 11.9% | 2365 | 26.0% | 3058 | 33.7% |
| General Surgery | 2465 | 9.1% | 559 | 22.7% | 302 | 12.3% | 653 | 26.5% | 252 | 10.2% | 170 | 6.9% | 324 | 13.1% | 886 | 35.9% |
| Accident and Emergency | 1768 | 6.5% | NA | NA | NA | NA | NA | NA | 288 | 16.3% | 198 | 11.2% | 354 | 20.0% | 361 | 20.4% |
| Gastroenterology | 982 | 3.6% | 336 | 34.2% | 151 | 15.4% | 379 | 38.6% | 336 | 34.2% | 151 | 15.4% | 379 | 38.6% | 379 | 38.6% |
| **Total Patients** | 27063 |  | 4967 | 18.4% | 1968 | 7.3% | 5673 | 21.0% | 4014 | 14.8% | 2412 | 8.9% | 4966 | 18.3% | 9035 | 33.4% |

Arranging follow up with gastroenterology was less common for patients admitted under ENT (12.2%) or General Surgery (13.1%), than other specialties (Accident and Emergency 20% and General Medicine 26%) (p<0.001).

### Potential impact of failure to take biopsies at endoscopy for foreign body in the oesophagus

In recent data from Scotland, 26% of patients with food bolus obstruction had EoE^15^. This suggests that in the present study cohort approximately 1300 patients missed an opportunity to be diagnosed with EoE, as oesophageal biopsies were not taken at endoscopy for FBO.

### Proposed algorithm for management of food bolus obstruction

Figure 2 shows a proposed algorithm for the management of patients presenting as an emergency with food bolus obstruction based on the study results and published literature.

**Figure 2.**
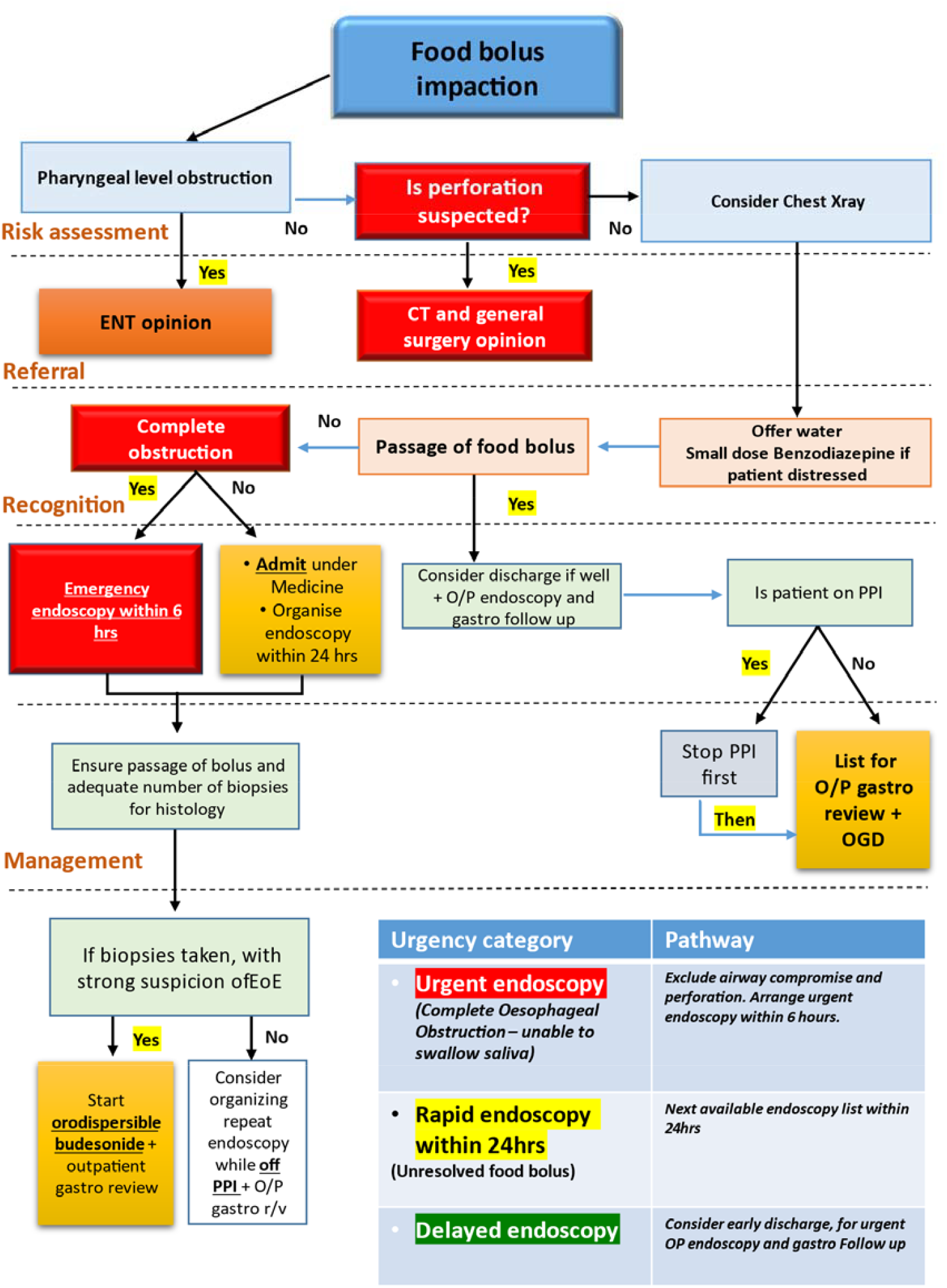
Proposed algorithm for the management of patients presenting as an emergency with food bolus obstruction.

## Discussion

Approximately 2500 patients present annually in England as an emergency with foreign body in the oesophagus. Over the study period, 75.7% of patients had an upper GI endoscopy but only 19.8% of patients had biopsies taken to exclude EoE and other pathology potentially responsible for food bolus obstruction. The rates of endoscopy with biopsy encouragingly increased over the study period from 13.2% in 2008 to 23.7% in 2019. Patients were less likely to undergo endoscopy with biopsy if they were admitted under ENT. 33.4% of patients with foreign body in the oesophagus were followed-up in outpatients in the 12 months following emergency admission with 18.3% being seen in gastroenterology outpatients.

The number of patients admitted to hospital with foreign body in the oesophagus increased by a third between 2008 and 2015. This is likely to reflect the increasing incidence of EoE that has been consistently reported in Europe and North America^16^. A Dutch cross-sectional study using 25 years of national histopathological data reported that the annual incidence of EoE increased from 0.01 per 100,000 of the population to 3.6 per 100,000^17^. The current annual incidence in the UK adult population is thought to be 7/100 000 people^18^.

As the number of patients admitted to hospital with foreign body in the oesophagus increased, the proportion undergoing endoscopy and biopsy also increased, particularly between 2015 and 2019. Patients were 80% more likely to undergo endoscopy with biopsy in 2018 compared with 2010 and awareness of EOE as a leading cause of foreign body obstruction is likely to have contributed to this improvement. The increase in biopsy rates in more recent years corresponds with publication of the European Society of Gastrointestinal Endoscopy clinical guideline for removal of foreign bodies from the upper GI tract in adults which advocates multilevel biopsy to exclude EOE in food bolus obstruction^8^. Despite the increase in endoscopy with biopsy over the study period, the biopsy rate remained much lower than it should be. In 2018, the year with the highest biopsy rate, only 25.3% of patients had biopsies. A US study of 548 patients with oesophageal foreign body impaction revealed similar findings with only 27% of patients who underwent endoscopy having oesophageal biopsies^4^. Failing to take biopsies to exclude EoE during endoscopy for food bolus obstruction represents an important missed opportunity to treat EoE to prevent further emergency admissions. From the prevalence of EoE in patients presenting with food bolus obstruction in a recent Scottish series, we estimate that an additional 1300 patients would have been diagnosed with EoE if biopsies had been taken as recommended^14,15^.

Patients admitted under ENT were much less likely to undergo endoscopy and endoscopy with biopsy compared to other specialties. This suggests a divergence in practice between specialties in managing foreign body obstruction and a lack of awareness of EoE as a cause of food bolus obstruction among ENT surgeons. Using rigid endoscopy for food bolus extraction rather than flexible endoscopy, given the difficulties taking biopsies with the former, is also likely to have contributed. Other studies have also reported lower biopsy and endoscopy rates in patients admitted under ENT and General Surgery^15^.

A similar pattern was observed in terms of patient follow-up after an episode of foreign body in the oesophagus. Overall, one third of patients received follow-up either with the same specialty or with gastroenterology. General Medicine referred 26% of its cohort of patients to gastroenterology compared to 12% from ENT and 13% from General Surgery. The lack of appropriate follow-up for such patients has been shown to be associated with recurrent food bolus impaction and complications^8,19,20^. Appropriate follow-up provides the opportunity to ensure a repeat endoscopy and biopsy is arranged if this was not undertaken during the acute admission with foreign body in the oesophagus.

0.9% of patients underwent a dilatation procedure during endoscopy and only 11.6% of these patients were coded as having biopsies taken. Endoscopic features such as Schatzki rings, furrows and strictures are associated with EoE, emphasising the importance of obtaining biopsies in such cases to exclude EoE and other aetiologies for stricturing and food bolus obstruction (ref EoE guidelines). The incidence of perforation during diagnostic upper GI endoscopy is 0.03% but this increases to between 0.09-2.2% with dilatation^21,22^ 0.4% of endoscopies in the study population were complicated by oesophageal perforation emphasising the importance of diagnosing and treating the underlying cause to prevent recurrent episodes of food bolus obstruction given these attendant risks.

This study has significant strengths in terms of patient numbers, national coverage and the reliability of procedural coding. However, HES is unable to provide detail related to the number or position of biopsies taken during endoscopy and does not include the histological results, so we are unable to draw conclusions as to the prevalence of EOE or compliance with recommended numbers and sites of biopsy. HES does not include detailed information on the results of endoscopy so we were unable to estimate how many patients had alternative causes for their foreign body in the oesophagus such as objects other than food or alternate causes of food bolus obstruction such as gastro-oesophageal reflux disease. In the validation exercise, 19% of patients coded as foreign body in the oesophagus had a foreign body other than just food. The number of patients undergoing both endoscopy and endoscopy with biopsy and outpatient follow-up following an episode of foreign body in the oesophagus needs to be interpreted in light of this. Patients deliberately swallowing foreign objects such as batteries may not require endoscopy and certainly would not require biopsies to exclude EOE^1^.

## Conclusions

The key finding from this study is that oesophageal biopsies to exclude EoE were only obtained during endoscopy for foreign body in the oesophagus in 20% of patients, despite 76% undergoing endoscopy. There were wide variations in endoscopic, biopsy and follow-up practices between different specialties, particularly for patients admitted under ENT. An algorithm for inpatient management of food bolus obstruction is proposed.

## Abbreviations

EOE: eosinophillic oesophagitis
OR: odds ratio
CI: confidence intervals
HES: Hospital Episode Statistics
IQR: interquartile range

## Author contributions

Study concept and design was jointly conceived by RA, DK and NT. Data extraction and analyses were performed by BC and DM. The manuscript was drafted by RA and NT. This was critically reviewed and approved by all authors.

## Data Availability statement

HES data are available under a data sharing agreement with NHS Digital for the purposes of service evaluation and are not available for open access.

